# Histological correlates of hippocampal magnetization transfer images in drug-resistant temporal lobe epilepsy patients

**DOI:** 10.1101/2020.07.10.20150953

**Authors:** Jose Eduardo Peixoto-Santos, Tonicarlo R Velasco, Carlos Gilberto Carlotti, Joao Alberto Assirati, Gustavo Henrique de Souza e Rezende, Katja Kobow, Roland Coras, Ingmar Blümcke, Carlos Ernesto Garrido Salmon, Antonio Carlos dos Santos, Joao Pereira Leite

## Abstract

**Objective:** Temporal lobe epilepsy patients (TLE) often present with hippocampal atrophy, increased T2 relaxation, and reduced magnetization transfer ratio (MTR) in magnetic resonance images (MRI). The histological correlates of the reduced MTR are so far unknown. Since MTR is dependent on the tissue’s macromolecules, our aim was to evaluate the correlations between cellular populations, extracellular matrix molecules and the MTR in TLE patients.

**Methods:** Patients with TLE (n=27) and voluntaries (=20) were scanned in a 3 Tesla MRI scanner, and MTR images were calculated from 3DT1 sequences with magnetization pulse *on resonance*. Immunohistochemistry for neurons, reactive astrocytes, activated microglia, and extracellular matrix chondroitin sulfate were performed in formalin fixed, paraffin embedded tissues of TLE and autopsy controls (n=10). Results were considered significant at p<0.05.

**Results:** Compared to the respective controls, TLE patients had reduced hippocampal MTR, increased reactive astrocytes and activated microglia, increased extracellular chondroitin sulfate, and reduced neuron density, compares to controls. MTR correlated positively with neuron density in CA3 and with chondroitin sulfate in CA3 and CA1. Multiple linear regressions reinforced the correlations between chondroitin sulfate and MTR.

**Significance:** Our data indicate that extracellular matrix molecules are the most significant histological correlates of magnetization transfer ratio in the hippocampus of TLE patients.

## 1. Introduction

Hippocampal sclerosis (HS) is the most common pathological finding in adults with drug-resistant epilepsy ^4^. In the pathological evaluation, HS is characterized by differential neuron loss, often more severe in CA1 and CA4 ^5^. Besides neuron loss, gliosis and changes in extracellular matrix proteins are seen in the sclerotic hippocampus of these patients ^8; 25; 26; 37^. Moreover, gliosis can even be seen in the hippocampi of temporal lobe epilepsy patients (TLE) without neuron loss ^34^. The presurgical magnetic resonance imaging (MRI) of these patients often shows hippocampal volume loss and increased T2 signal ^2; 6; 7^. However, in up to 15 % of the TLE patients, no abnormalities are observed in the standard MR evaluation ^3; 17^. Although the normal MRI patients often have only milder or no neuron loss, some can have neuron loss and other pathological changes as severe as those cases with hippocampal sclerosis detected on MRI ^17; 25^. Since the presence of HS is often an indicator of good surgical outcome in TLE ^5; 34^, it is crucial for improving the pre-surgical detection of those MRI-negative cases with pathologically proven HS, as well as more subtle pathological changes in the hippocampus.

New quantitative MRI techniques could improve the detection of more subtle abnormalities seen in epilepsy patients ^11^. Magnetic transfer (MT), an imaging protocol that evaluates macromolecules not visible in either T1-weighted or T2-weighted images, is often reduced in regions with EEG abnormalities with normal conventional MRI ^29^. While some studies have indicated the association between myelin loss and MTR reduction in the white matter, and between neuron loss and MTR reduction in the cerebral cortex, there remains to be determined which pathological changes are responsible for the decreased hippocampal MTR in TLE patients. We recently found that extracellular matrix chondroitin sulfate proteoglycan (CSPG) could impact both hippocampal volume ^25^ and T2 signal relaxation time ^24^. Since MTR is influenced by the macromolecules present in the tissue, our objective was to evaluate the correlations between the MTR and chondroitin sulfate, as well as with cellular populations, in the hippocampus of drug-resistant temporal lobe epilepsy patients.

## 2. Materials and Methods

### 2.1. Patients

Twenty-six drug-resistant TLE patients were selected during the presurgical evaluation at the Epilepsy Surgery Centre (CIREP) of Ribeirao Preto Medical School. Presurgical workup included history review, neurological evaluation, neuropsychiatric memory tests, video-EEG, and an optimized MRI protocol for TLE, for the definition of epileptogenic focus. Patients with undoubted seizure focus underwent standard *en bloc* temporal lobe resection.

Thirty age-matched control cases consisted on: twenty healthy volunteers that underwent the same presurgical MRI protocol used for TLE cases (radiological controls, RC), used for defining MRI differences in TLE; ten autopsy cases (histological controls, HC) whose hippocampi were collected to serve as a standard for immunohistochemistry analysis.

Inclusion criteria were: age between 20 and 60 years for all groups; diagnosis of drug-resistant TLE (for TLE group); quantitative MRI protocol (for TLE and RC). Exclusion criteria were: the presence of MRI abnormalities (for RC); generalized or extratemporal EEG spikes; the presence of pathological changes in the hippocampus in histological evaluation (for HC); the presence of other brain pathology than hippocampal sclerosis (for TLE cases); postmortem time superior to 12 hours (for HC).

This study was registered in the Brazilian’s Health Ministry and approved by the local Research Ethics Committee of the Hospital das Clinicas (HCRP # 7200/2016). A written Informed Consent Term, previously approved by the Research Ethics Committee, was obtained from all patients or next-of-kin enrolled in this study, following the Declaration of Helsinki.

### 2.2. MRI Protocol

All TLE cases and RC volunteers underwent MRI in a Philips Achieva 3.0T X-series with an 8 elements phase-array head coil. For the definition of hippocampal atrophy, 3D single shot T1-weighted images were performed (TE = 3.2 ms; TR = 7 ms; flip angle = 8°; inversion pulse = 900 ms; shot interval = 2500 ms; voxel size = 1 mm^3^; FOV = 240×240 mm; acquisition time = 4.5 minutes). T2 relaxation was additionally measured with 2D turbo spin echo sequences (TEs = 20, 40, 60, 80, 100 ms; TR = 3000 ms; flip angle = 90°; EPI factor = 5; voxel size = 1×1×3 mm; FOV = 240×180 mm; acquisition time = 4 minutes). Magnetization transfer ratio was evaluated with two 3D sequences with minimal T1 and T2 weighting (TFE = 3; TE = 3 ms; TR = 3.6 ms; flip angle = 8°; magnetization transfer saturation pulse *on resonance*; voxel = 1×1×3 mm; FOV = 240×180, acquisition time = 5 minutes).

The evaluation of hippocampal volume, T2 relaxation, and magnetization transfer ratio was performed with MINC Tools (BIC, McGill, Canada) and homemade scripts for the calculation of relaxation and MTR maps.

### 2.3. Immunohistochemistry Protocol

Coronal sections from the hippocampal body of TLE and HC cases were fixed in formalin, dehydrated, clarified, and embedded in paraffin. Eight-micrometer-thick sections were submitted to immunohistochemistry with anti-NeuN (#MAB377, Chemicon; a neuronal marker), anti-GFAP (#M0761, Dako; a marker of reactive astrocytes), anti-HLA-DR (#M0746, Dako; a marker of activated microglia), and anti-CSPG (#C8035, Sigma; a broad-spectrum chondroitin sulfate proteoglycan marker), following protocols and dilutions previously published ^24^.

Micrographs from the regions of interest were collected with an AxioCamMR5 in an Axio Imager M1 microscope with the AxioVision 4.8.1 software (Zeiss). Illumination was maintained constant (3V), and exposure varied from each protein (33 ms for NeuN and GFAP, 60 ms for HLA-DR, and 40 ms for CSPG), and micrographs were taken at 100x magnification for NeuN, GFAP, and CSPG, and at 200x magnification for HLA-DR. The analysis was performed with ImageJ 1.45s software (NIH), with a semi-quantitative analysis of the immunopositive area fraction ^13; 18; 24^. The thresholds used were 154 ± 10 for GFAP, 50 ± 10 for HLA-DR, and 132 for CSPG. Neuron density was estimated following Abercrombie’s method, as described elsewhere ^23^. The TLE cases were classified according to the hippocampal sclerosis type ^5^. The regions of interest where the hippocampal subfields CA4, CA3, CA2, CA1, and subiculum, as delineated by ILAE Taskforce ^5^.

### 2.4. Statistics

For the parametric variables, ANOVA with Bonferroni *post hoc* test or Student’s t-test were performed, whereas Kruskal-Wallis with Dunn *post hoc* test or Mann-Whitney’s test were used for non-parametric data. Spearman’s correlation was performed to evaluate the association between histological and MR data, and multiple linear models were constructed based on the most relevant results. The association between categorical data and MTR class was explored with logistic regression followed by a χ^2^ test. Results were considered significant at p < 0.05.

## 3. Results

### 3.1. Clinical Data

All subject enrolled in the present study were age-matched (HC = 46.9 ± 13.6 years, RC = 43.6 ± 7.6 years, and TLE = 43.2 ± 11.8 years; p = 0.635). Postmortem interval was of 8.3 ± 4.1 hours, and most HC cases had heart failure as the main cause of death (50 %), followed by sepsis (30 %) and pneumonia (20 %). TLE cases had the first seizure at 10.1 ± 10.0 years (median of 5.5 years, ranging from 0.5 to 35 years) and seizure recurrence at 18.0 ± 11.1 years (median of 17 years, ranging from 0.5 to 45 years). All remaining clinical data are described in **Table 1**.

**Table 1.**
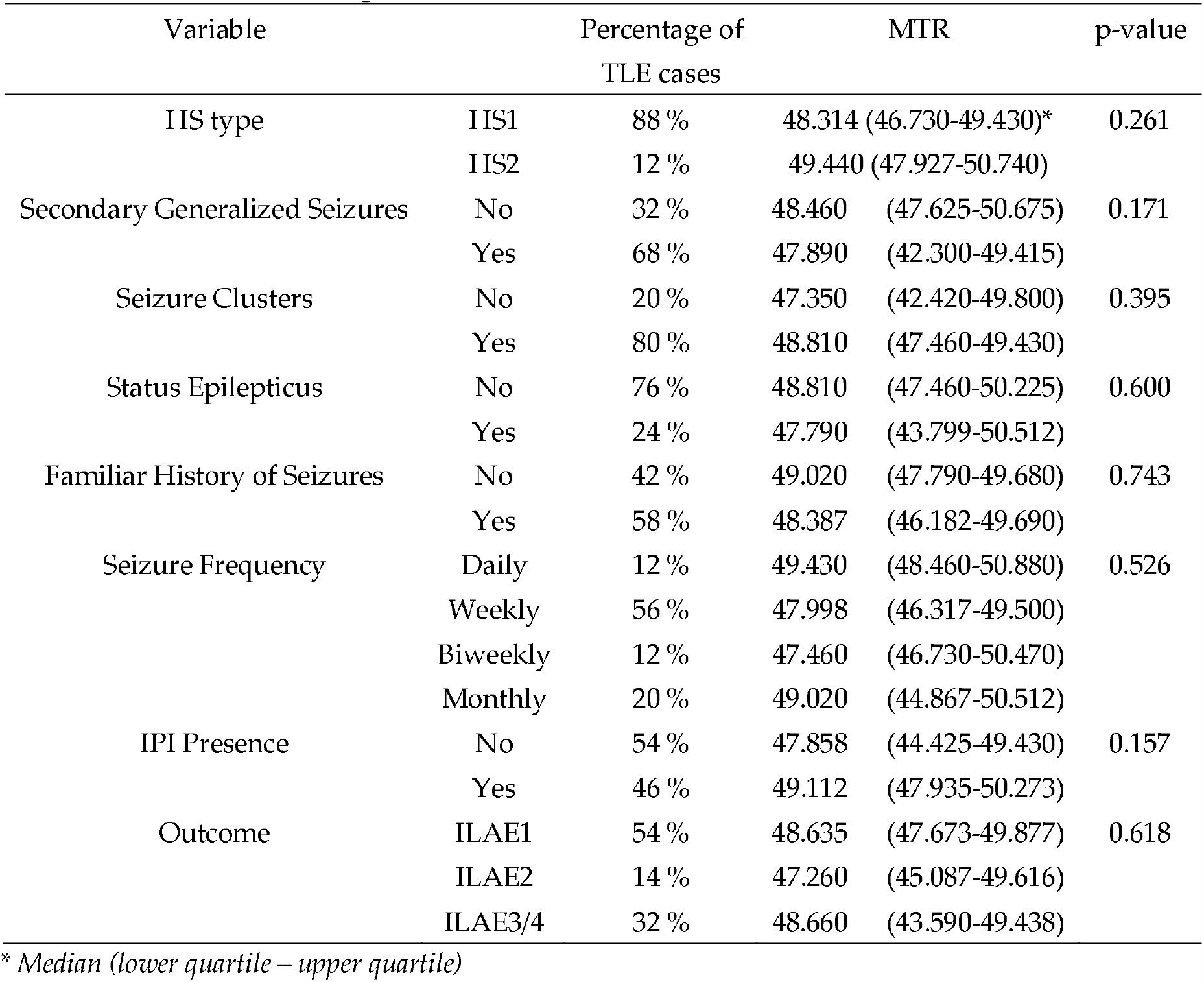
MTR value according to clinical data classes. al hippocampal MTR are shown in green.

| Variable |  | Percentage of<br>TLE cases | MTR | p-value |
| --- | --- | --- | --- | --- |
| HS type | HS1 | 88 % | 48.314 (46.730-49.430)* | 0.261 |
|  | HS2 | 12 % | 49.440 (47.927-50.740) |  |
| Secondary Generalized Seizures | No | 32 % | 48.460 (47.625-50.675) | 0.171 |
|  | Yes | 68 % | 47.890 (42.300-49.415) |  |
| Seizure Clusters | No | 20 % | 47.350 (42.420-49.800) | 0.395 |
|  | Yes | 80 % | 48.810 (47.460-49.430) |  |
| Status Epilepticus | No | 76 % | 48.810 (47.460-50.225) | 0.600 |
|  | Yes | 24 % | 47.790 (43.799-50.512) |  |
| Familiar History of Seizures | No | 42 % | 49.020 (47.790-49.680) | 0.743 |
|  | Yes | 58 % | 48.387 (46.182-49.690) |  |
| Seizure Frequency | Daily | 12 % | 49.430 (48.460-50.880) | 0.526 |
|  | Weekly | 56 % | 47.998 (46.317-49.500) |  |
|  | Biweekly | 12 % | 47.460 (46.730-50.470) |  |
|  | Monthly | 20 % | 49.020 (44.867-50.512) |  |
| IPI Presence | No | 54 % | 47.858 (44.425-49.430) | 0.157 |
|  | Yes | 46 % | 49.112 (47.935-50.273) |  |
| Outcome | ILAE1 | 54 % | 48.635 (47.673-49.877) | 0.618 |
|  | ILAE2 | 14 % | 47.260 (45.087-49.616) |  |
|  | ILAE3/4 | 32 % | 48.660 (43.590-49.438) |  |

\* Median (lower quartile – upper quartile)

### 3.2. MRI Evaluation

The automatic hippocampal analysis showed no significant difference in hippocampal volume between controls and TLE cases (p = 0.104; **Figure 1 A**). Only nine of the TLE patients (34.6 %) had hippocampal atrophy, following the cutoffs of previous quantitative studies with a different set of cases ^25^. Hippocampus from TLE patients presented with higher relaxation time than controls (p < 0.001; **Figure 1 B**). The hippocampi from TLE cases had a significant reduction in MTR compared to controls (p < 0.001; **Figure 1 C**). Based on control hippocampi average and standard deviation, cases below 95% MTR of controls (i.e., below 48.6 % magnetization transfer) were classified as having low MTR, and those above as having normal MTR. In the patients without hippocampal volume loss, the low MTR was seen in 50 % of cases. As for the cases with hippocampal atrophy, 88 % had low MTR. The combination with T2 relaxation increases the detection of the abnormal hippocampus to 64 % of normal volume cases.

**Figure 1.**
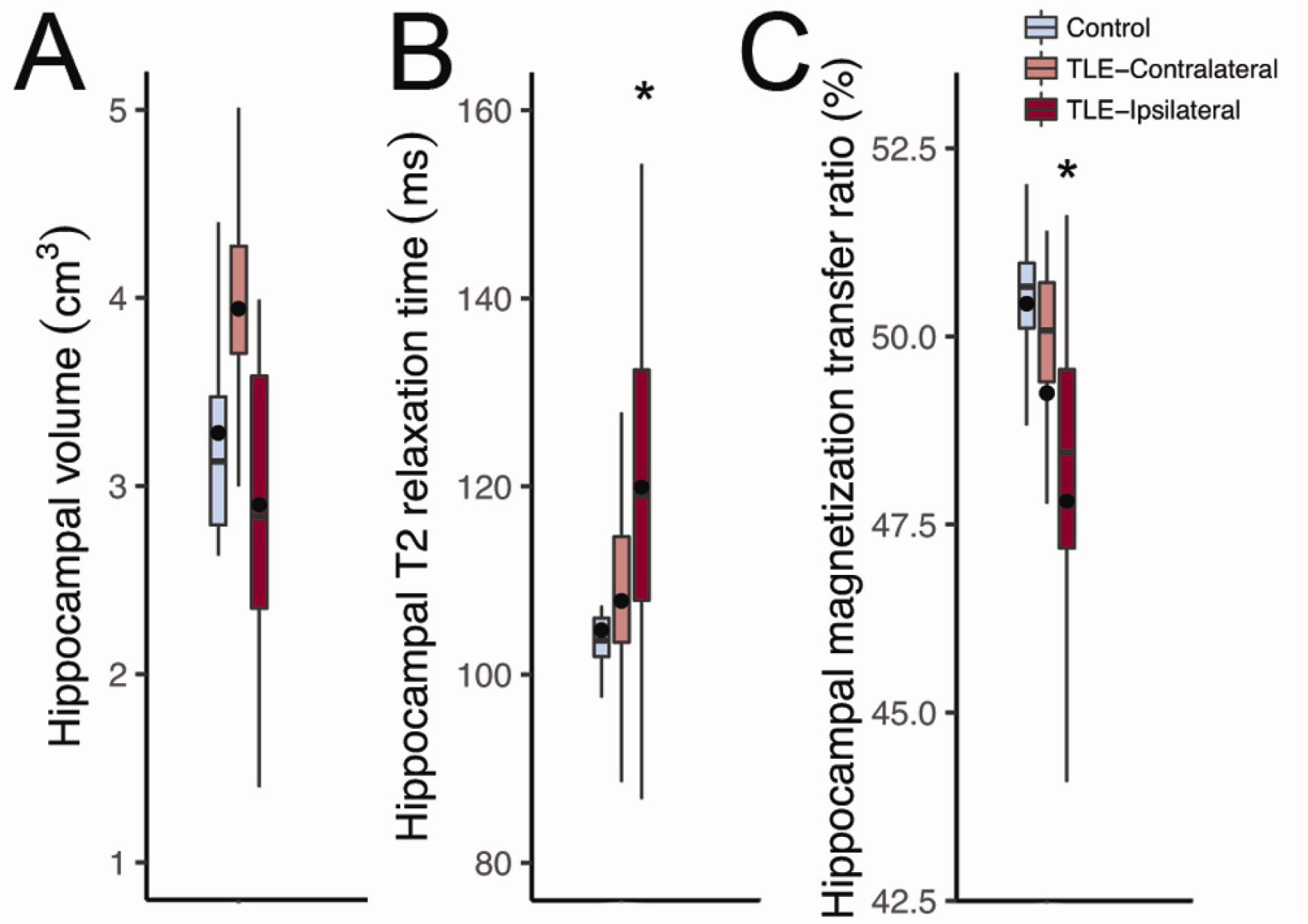
Quantitative magnetic resonance evaluation of the hippocampi from controls (radiological controls, blue boxplots) and TLE patients (red boxplots). (A) There was no difference between controls and ipsilateral (dark red boxplot) or contralateral (light red boxplot) hippocampi from TLE regarding volume. (B) Only the ipsilateral hippocampus of TLE patients presented with increased T2 relaxation time and (C) reduced magnetization transfer, when compared to controls. There was no difference between the ipsilateral and contralateral hippocampus of TLE cases. The asterisks indicate statistical difference, the line inside the boxplots indicate median and the dot indicate mean.

### 3.3. Cellular Populations and Extracellular Matrix

TLE patients had lower neuron density than controls in CA4, CA3, CA2, and CA1 (p < 0.001), with no difference in the subiculum (p = 0.57; **Figure 2 A**). Reactive astrogliosis was seen in CA1 (p = 0.001) and in the subiculum (p = 0.002; **Figure 2 B**) of TLE cases, compared to controls. Activated microglia was present in CA4 (p = 0.012), CA3 (p = 0.004), CA2 (p = 0.003), and in CA1 (p < 0.001; **Fig. 2 C**) of TLE patients, compared to controls. Increased expression of chondroitin sulfate proteoglycan was seen in all hippocampal subfields evaluated (p < 0.001; **Figure 2 D**). A representative image showing the expression patterns in TLE is shown in **Figure 3**.

**Figure 2.**
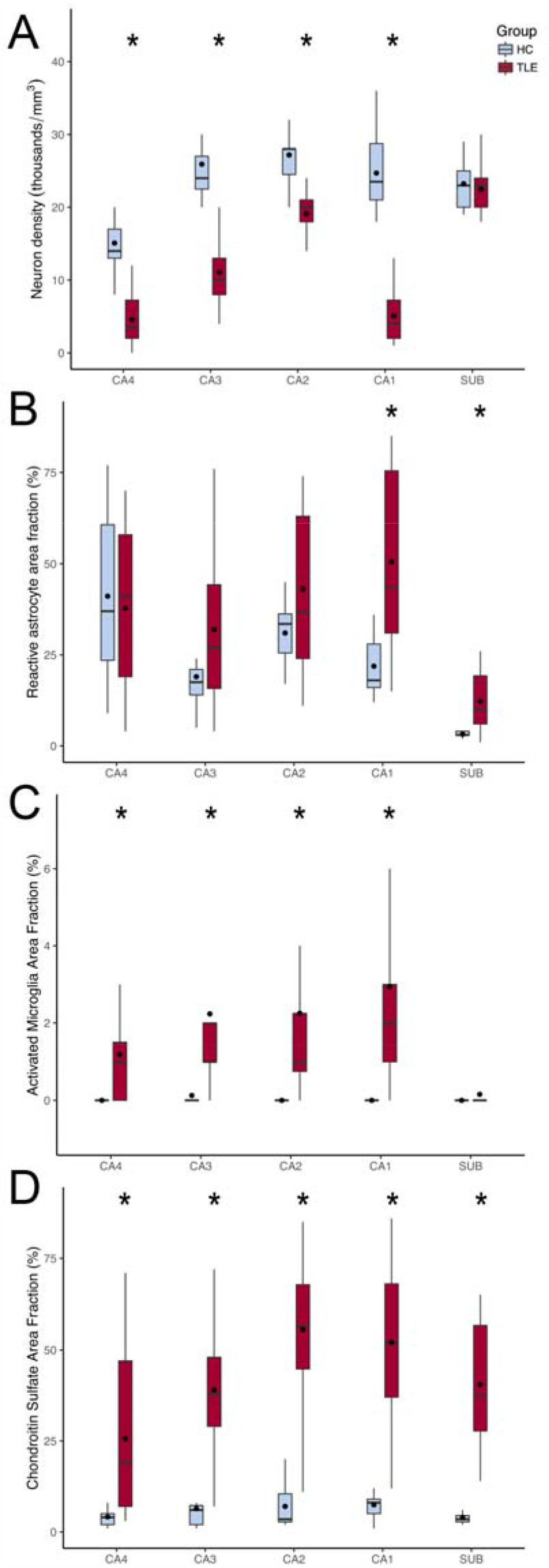
Semi-quantitative evaluation of hippocampal sections from controls (histological controls, blue boxplots) and TLE patients (red boxplots) submitted to immunohistochemistry. (A) All hippocampal subfields but the subiculum of TLE cases presented with neuron loss. (B) Only CA1 and the subiculum of TLE had significant astroglial reaction, when compared to controls. (C) Following the neuron density changes, all hippocampal subfields but the subiculum had activated microglia, when compared to controls. (D) Chondroitin sulfate proteoglycan was seen in higher levels in all hippocampal subfields of TLE cases, compared to controls. The asterisks indicate statistical difference, the line inside the boxplots indicate median and the dot indicate mean.

**Figure 3.**
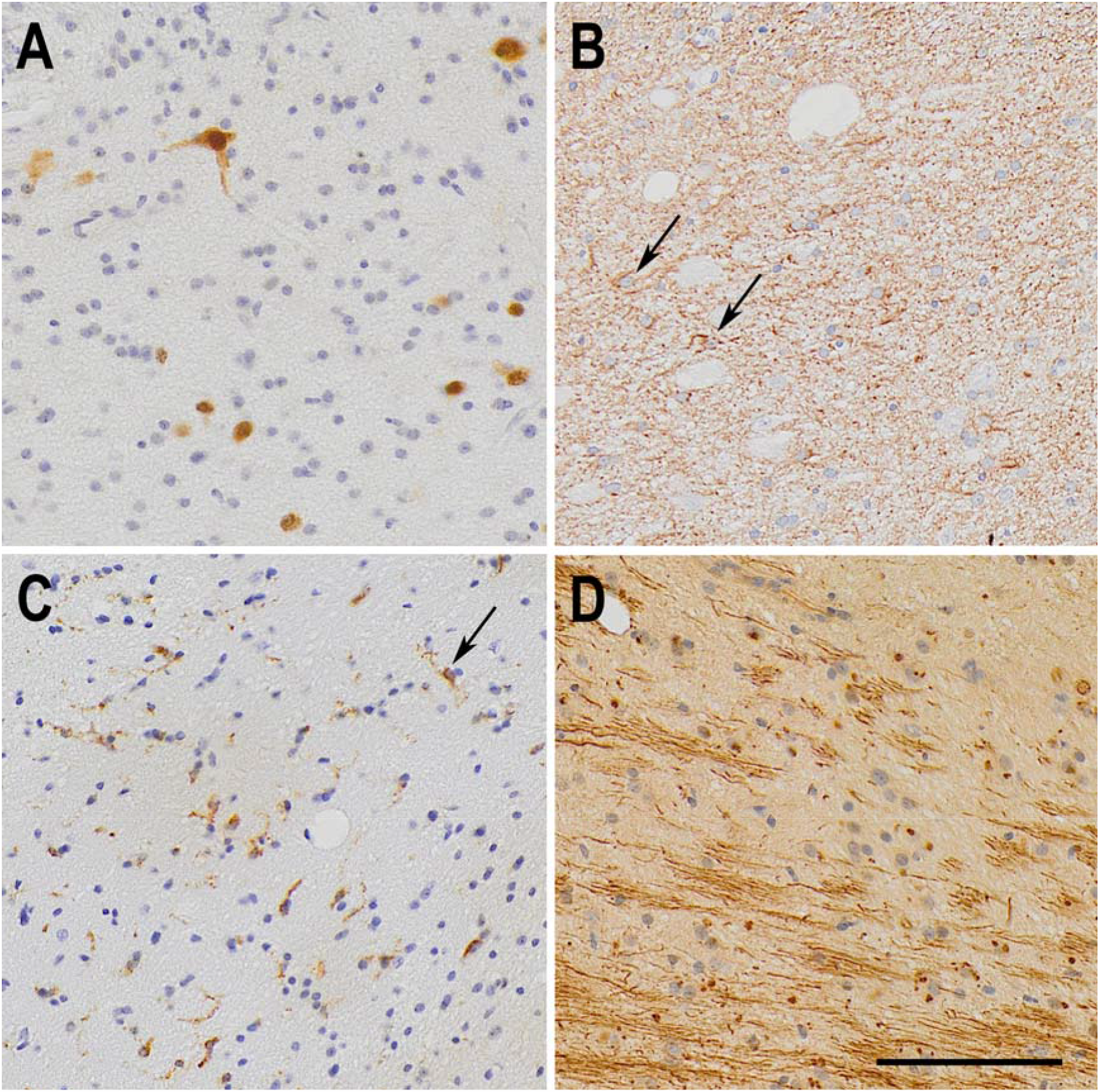
Representative immunohistochemistry micrographs of the pathological changes seen in the CA1 subfield from TLE samples. (A) NeuN staining, showing reduced neuron density in TLE CA1. (B) GFAP staining showing fibrous astrogliosis, as defined by ILAE, in a TLE case. Two reactive astrocytes are indicated by arrows. (C) Increased activation of microglial cells, marked by the expression of MHC class II HLA-DR protein. One activated microglia is pointed by the black arrow. (D) Fibrous aggregates of extracellular chondroitin sulfate proteoglycans, showed by CS-56 antigen immunohistochemistry. The bar in (D) indicates 100 µm.

### 3.4. Associations between MRI, clinical data, and histology

Hippocampal MTR were not significantly different regarding HS type, the presence of secondarily generalized seizures, the occurrence of status epilepticus or seizures in clusters, a positive familial history of seizures, seizure frequency, occurrence of IPI, or surgical outcome (see **Table 1**). Logistic regression with the MTR classification (low vs. normal) reinforced the lack of association between MTR and the clinical data.

Hippocampal MTR correlated positively with neuron density in CA3 (R = 0.581, p = 0.0059; **Figure 4 A**) and with CSPG levels in CA4 (R = 0.513, p = 0.0291), CA3 (R = 0.66, p = 0.0038; **Figure 4 B**), CA2 (R = 0.486, p = 0.0297), CA1 (R = 0.544, p = 0.0109; **Figure 4 C**), and in the subiculum (R = 0.578, p = 0.0182; **Figure 4 D**). Negative correlations between hippocampal MTR and activated microglia were seen in CA3 (R = −0.566, p = 0.0175), CA2 (R = −0.636, p = 0.0079), and in CA1 (R = - 0.604, p = 0.0103). Reactive astrocytes had no correlation with MTR. A 3D visualization of neuron density, chondroitin sulfate, and MTR can be seen, respectively, in **Supplementary Figures 1** and **2**.

**Figure 4.**
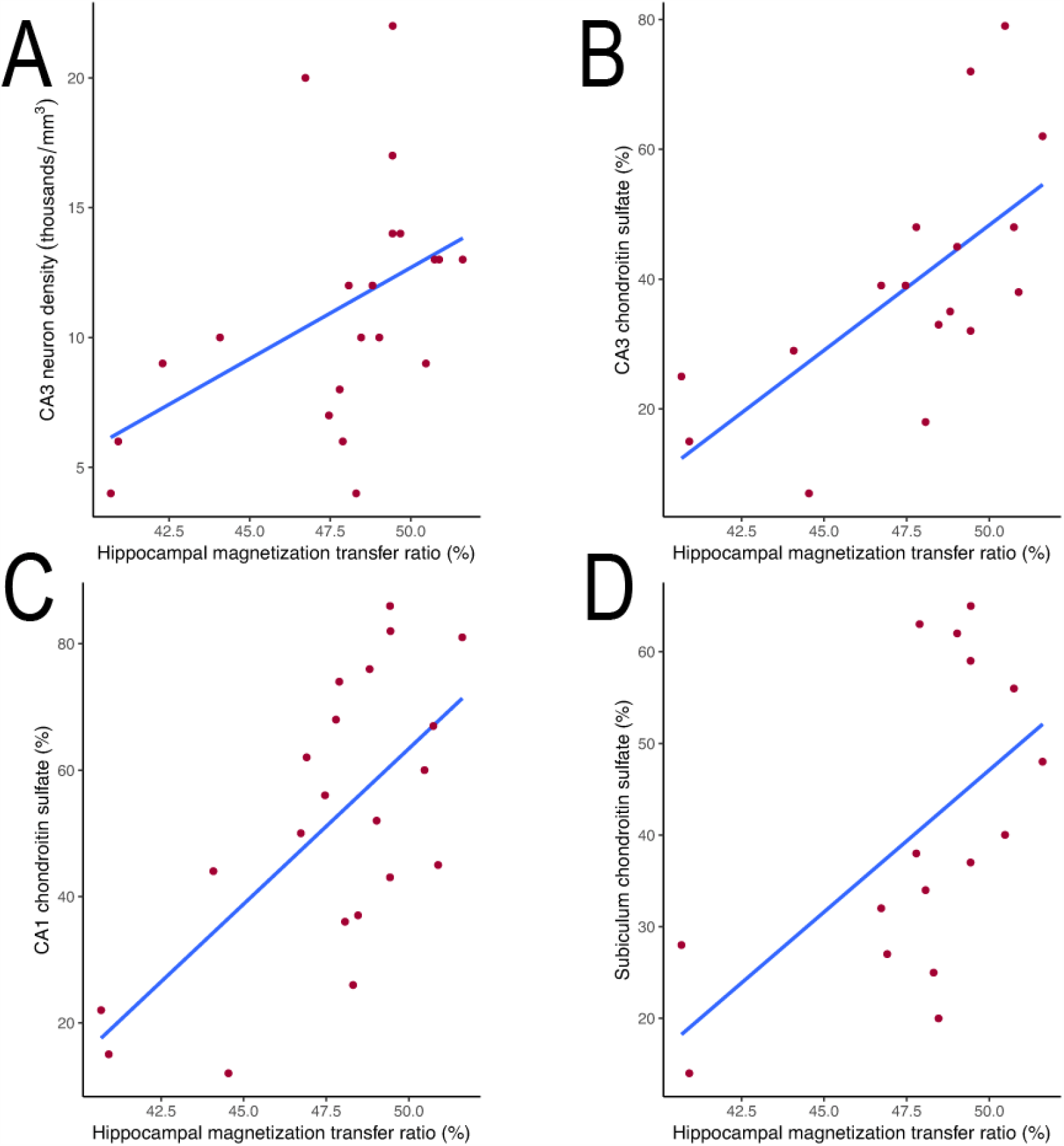
Scatterplot and regression between histological and magnetization transfer data of TLE patients. Whole hippocampal MTR from TLE cases correlated positively with neuron density in CA3 (A) and with chondroitin sulfate levels in CA3 (B), CA1 (C), and the subiculum (D).

From different combinations of neuron density and CSPG levels in CA4, CA3, CA1, and the subiculum, the best multiple linear model for explaining the variation in hippocampal MTR were the combination of neuron density and CSPG in CA3 and CA1, with a overall predictive value of 71 % for the MTR (R = 0.841, R^2^ = 0.708, VIF < 2.971, p = 0.006). The following equation was given for these variables:

MTR = 40.282 + (0.324 × neuron density in CA3) - (0.274 × neuron density in CA1) + (0.0750 × CSPG levels in CA3) + (0.0429 × CSPG levels in CA1)

Individually, the changes in CA3 explain 59 % of the hippocampal MTR (R = 0.767, R^2^ = 0.588, VIF = 1.049 p = 0.003), whereas CA1 explains 43 % of MTR (R = 0.652, R^2^ = 0.426, VIF = 1.035, p = 0.009). The multiple linear regression equations are as follows:

MTR = 39.779 + (0.349 × neuron density in CA3) + (0.100 × CSPG levels in CA3) MTR = 43.487 - (0.131 × neuron density in CA1) + (0.0943 × CSPG levels in CA1)

## 4. Discussion

Reduced hippocampal volume and increased T2-weighted signal are important hallmarks of the epileptogenic hippocampi in drug-resistant TLE patients ^2; 6; 7^. However, some patients have no MRI abnormalities in the standard MRI sequences ^3; 17^. In these cases, new MRI techniques, such as MTR, could help improve the MRI detection of the epileptogenic focus. Our present study evaluated MTR in TLE patients with different hippocampal volumes on MRI, showing reduced MTR in the epileptogenic hippocampi. Moreover, in patients whose hippocampus showed normal volume, MTR were below normal values in 50 %, thus indicating that in normal volume cases MTR could help define the epileptogenic hippocampus, and in two cases (8 % of TLE) only MTR was below normal levels. This finding agrees with previous data in which reduced MTR was seen in the hippocampus associated with seizure focus ^36^, and indicates that MTR can help lateralize the focus in some cases.

The atrophy seen in the epileptogenic hippocampus was linked to the degree of neuron loss in the granule cell layer ^6^ but more consistently to neuron loss in the CA1 subfield ^10; 37^. A previous study from our group also pointed out neuron loss in CA1, together with chondroitin sulfate levels in this subfield, as the most important factor for explaining hippocampal volume ^25^. Reduced magnetization transfer has been linked with myelin and neuron (axonal) loss in multiple sclerosis ^14; 22; 38^. An animal model of autoimmune encephalomyelitis further indicated that MTR reduction in regions of neuron loss is more evident than in those with only demyelination ^27^. In TLE, neuron loss is believed to be associated with MTR reduction ^15^. However, a study with temporal cortex showed no correlation between neuronal population and MTR values ^13^. Our data indicate that neuron loss in CA3 is important for MTR reduction seen in TLE.

From the glial population evaluated with immunohistochemistry, only the activated microglia correlated with the hippocampal MTR. The glial reaction is a common finding in the hippocampus of drug-resistant TLE patients ^18; 24; 25^. Gliosis often following the degree of neuronal loss and seizure severity ^8; 19^. However, the glial reaction is known to occur in the absence of neuron loss ^34^. Increased T2 signal and relaxation time is often linked to reactive astrogliosis ^6; 37^, whereas no MRI sequence so far has been directly linked to microglia ^1; 12; 20^. We saw negative correlations between MTR and activated microglia, but not with reactive astrocytes. Given the usually low increase in activated microglia in the hippocampus when compared to the often exuberant neuron loss and astroglial reaction, it would be unexpected for microglia to affect MRI sequences. It is possible that the inverse correlation between microglia and MTR is an indirect effect from the stronger association between neuron loss and CSPG with MTR. In fact, when added to the multiple linear regression model that included CA3 and CA1 values, the activated microglia presents the highest VIF values (2.935 for CA3 and 4.037 for CA1), indicating multicollinearity between microglia and other factors. Thus, we believe activated microglia should be seen as an indirect indicator for MTR changes.

Extracellular matrix in the central nervous system is often studied concerning tissue plasticity ^16; 32^. Increased CSPG levels were already described in patients with epilepsy and animal models ^26; 30^. In the pathology/MRI correlation field, some studies have shown an association between apparent diffusion coefficient changes and increased levels of CSPG ^28; 39^. Given that extracellular matrix accounts for 20 % of the brain parenchyma ^9; 33^, it is expected that these molecules are important for MRI changes. In fact, previous studies of our group with hippocampi of TLE patients have shown a significant association between CSPG and changes in hippocampal volume and T2 relaxation ^24; 25^. The link between extracellular matrix molecules and magnetization transfer is underexplored, and studies have provided mixed results. In phantoms of collagen and CSPG, while a study showed no correlation CSPG and MTR ^31^, another one has shown a weak positive association between CSPG and MTR ^21^. Studies with animal cartilage also indicated that MTR is more often affected by the amount of collagen, with little effect of CSPG ^21; 35^. Our present data indicate that hippocampal MTR is strongly associated with the levels of CSPG. Moreover, our 3D scatterplots allow the visualization of a complex scenario where some patients with normal MTR have both higher neuron density and CSPG content, whereas other have high CSPG and low neuron density or, less frequently, low CSPG and higher neuron density (see **Supplementary Figures 1 and 2**). In a previous study, we also saw similar indications that some patients with severe neuron loss can have a high content of CSPG and, thus, a normal hippocampal volume ^25^. In summary, the presence of normal hippocampal MTR should not rule out the occurrence of hippocampal neuron loss and, thus, hippocampal sclerosis.

The most important limitation of the present study is, in our view, that we only evaluated extracellular matrix with a broad-spectrum anti-chondroitin sulfate antibody, when it would be important to evaluate CSPG subtypes that also have important effects on tissue excitability, such as phosphacan (the molecules that compose the perineuronal nets) and neurocan, as well as hyaluronic acid. However, these molecules are better evaluated with western blot, which was not feasible for our samples. Moreover, it would be interesting to investigate larger series, especially with more patients with HS type 2, to evaluate the possible importance of MTR in separating HS type 1 and 2. These HS types are usually not distinguishable in presurgical MRI but can be identified in high-field ex-vivo MRI with smaller voxel size ^23^. Even if more HS type 1 cases where added, we believe stronger associations would emerge. Another critical problem of the MRI-histology correlational studies that is often not discussed is the problem of associating variables of different dimensionalities (i.e., one-mm-thick MRI signal of whole hippocampal data with eight-µm-thin slices of immunohistochemistry evaluated in ROIs of different subfields). If replaced by whole-hippocampus western blot, we would lose subfields variability but the general picture would be closer to whole hippocampal MTR values. The problem of comparing variables of different dimensions, as far as we know, has no clear, widely accepted solution. A better approach for future studies could be through immunohistochemistry evaluation of the hippocampal subfields on several slices throughout the hippocampus long axis and compare it with discrete MTR values of each subfield, and enrich the data with molecular approaches such as western blot and proteomics. Higher-field MRI with smaller voxels could also improve correlational studies.

## 5. Conclusions

The present study indicates that neuron density and extracellular matrix chondroitin sulfate are associated with MTR changes in the hippocampi of TLE patients, and together can explain more than 70% of the MTR signal in the ipsilateral hippocampus.

## Data Availability

Data from this study will be made available upon request

## Key Points

. In 64 % of patients with normal hippocampal volume on MRI, the combination of reduced magnetization transfer ratio and increased T2 relaxation indicates the epileptogenic hippocampus

. Extracellular matrix chondroitin sulfate proteoglycans are the most relevant correlate to magnetization transfer ratio in hippocampal sclerosis

. Increased chondroitin sulfate in the hippocampus can counterbalance the effect of neuron loss over the reduction in hippocampal magnetization transfer

## Author Contributions

conceptualization, J.E.P.S., C.E.G.S., A.C.S., J.P.L.; formal analysis, J.E.P.S., G.H.S.R., T.R.V; investigation, J.E.P.S., G.H.S.R.; resources, C.G.C., J.A.A., K.K., R.C., I.B., C.E.G.S., A.C.S., J.P.L.; writing—original draft preparation, J.E.P.S.; writing—review and editing, J.E.P.S., J.P.L.; project administration, J.P.L.; funding acquisition, J.E.P.S, J.P.S.

## Acknowledgments

We would like to thank the excellent technical support of Antonio Renato Meirelles e Silva, Geraldo Cassio dos Reis, and the staff from the CIREP, Magnetic Resonance Unit, Center of Imaging Sciences and Medical Physics (CCIFM), and Pathology Department.

## Conflicts of Interest

The authors declare no conflict of interest.

We confirm that we have read the Journal’s position on issues involved in ethical publication and affirm that this report is consistent with those guidelines.

## Abbreviations

ANOVA: Analysis of variance
CA: Cornu Ammonis
CSPG: Chondroitin sulfate proteoglycan
GFAP: Glial fibrilary acid protein
HLA-DR: Human leukocyte antigen - DR isotype
HR: Histological control
HS: Hippocampal sclerosis
ILAE: International League Against Epilepsy
IPI: Initial precipitating injury
MRI: Magnetic resonance imaging
MT: Magnetization transfer
MTR: Magnetization transfer ratio
NeuN: Neuronal nuclei
NIH: National Institutes of Health
RC: Radiological control
TLE: Temporal lobe epilepsy
VIF: Variance inflation factor

---

**Supplementary Figure 1**. 3D scatterplot showing the relations between hippocampal MTR and CA3 neuron density and chondroitin sulfate levels. Patients with MTR below normal values (low MTR) are shown in purple, whereas those with normal hippocampal MTR are shown in green.

**Supplementary Figure 2**. 3D scatterplot showing the relations between hippocampal MTR and CA1 neuron density and chondroitin sulfate levels. Patients with MTR below normal values (low MTR) are shown in purple, whereas those with norm

